# A Functional Sensor-to-Segment Orientation Method to Reduce the Effects of Varied Sensor Placement: Implications for IMU Best Practices

**DOI:** 10.1101/2022.11.29.22282894

**Authors:** Julien A. Mihy, Mayumi Wagatsuma, Stephen M. Cain, Jocelyn F. Hafer

## Abstract

To collect reliable data, it is important to determine how inertial measurement unit (IMU) sensor placement affects measurements of segment motion and to establish best practices for sensor placement and orientation procedures. We aimed to determine the extent to which a functional orientation method minimizes the effect of variations in sensor placement on IMU-derived segment excursions. Twenty healthy adults walked overground in a lab. Three IMUs were placed per segment on the pelvis, thigh, shank, and foot. Differences in estimated segment angular excursions between sensor placements were compared between an assumed sensor-to-segment orientation and two versions of a walking-based functional sensor-to-segment orientation. For time series data, differences in angular excursion between sensor placements were reduced from 60% of the gait cycle (assumed orientation) to 44% of the gait cycle (functional orientation) for the pelvis; from 31% (assumed) to 28% (functional) of the gait cycle for the thigh; and from 84% (assumed) to 0% (functional) of the gait cycle for the shank. Mean angular excursion RMSDs between sensor placements were <5° for most comparisons for the functionally oriented data. Functional orientation did not minimize inter-sensor placement effects when one sensor was located on a region with high soft tissue artifact (e.g., anterior thigh). Functional orientation reduced inter-sensor differences in segment excursion for the shank and foot and between-subject variance in inter-sensor differences for all segments. Functional orientation minimizes the effect of variations in IMU sensor placement, but care should be taken to select sensor placements that minimize soft-tissue artifact.

## Introduction

The use of inertial measurement units (IMUs) for gait research has increased in the last decade, largely because IMUs allow for affordable and portable measurement of movement. As IMUs are relatively new in biomechanics research, recommendations or standards for sensor placement for tracking body segment motion are lacking. Whether different sensor placements or small inter-individual or inter-session variations in sensor placement lead to meaningful differences in biomechanical outcomes is unknown. The use of IMUs in unobserved or real-world settings adds additional challenges if outcomes are affected by variations in sensor positioning within or between sessions. Functional sensor-to-segment orientation methods provide a potential means for capturing IMU data in a reliable reference frame and reducing differences due to sensor placement. However, the extent to which functional sensor-to-segment orientation minimizes the effects of variations in sensor placement has not been examined.

Several methods exist for orienting IMU data to interpretable (e.g., anatomical or functional) reference frames (Pacher et al., 2020; Vitali and Perkins, 2020). Segment reference frames can be created using various methods, the most common of which are assumed, functional, and model-based sensor-to-segment orientation methods. Assumed sensor-to-segment orientation, which relies on an IMU’s hardware axes being physically aligned with anatomy when placed on a body segment, requires no postprocessing to reorient IMU data. However, the accuracy of axis orientation in this method is sensitive to sensor placement and reliant on the assumption that sensors on the body surface align with underlying anatomy (Dejnabadi et al., 2005; Picerno et al., 2008; Sun et al., 2017). Functional sensor-to-segment orientation uses participant motion to orient IMU data to specific axes (e.g., approximately frontal, sagittal, or transverse) (Cain et al., 2016; Favre et al., 2009; Vargas-Valencia et al., 2016).

Functional orientation only requires simple motions to implement, but the orientations of the axes depend on the participant’s performance of those motions. Model-based methods use kinematic or statistical models of lower extremity degrees of freedom to orient data to anatomical reference frames. While model-based orientation minimizes reliance on precise sensor placement, these methods are computationally intensive, rely on physical and statistical assumptions of human movement, and may require extensive training data (McGrath and Stirling, 2020; Seel et al., 2014, 2012; Weygers et al., 2021). Because of assumptions about sensor orientation for assumed orientation and additional data or computation requirements for model-based orientation, functional orientation methods appear best-suited to most data collection scenarios, both in and out of the lab.

Functional sensor-to-segment orientation can be applied in several ways. Participants may perform a series of controlled segment rotations to orient IMU data to sagittal, frontal, and transverse anatomical axes (Favre et al., 2009, 2008; Seel et al., 2014). Accurate axis orientation thus relies on a participant’s ability to precisely perform controlled motions about each axis. Alternatively, a combination of static and dynamic data may be used to define functionally relevant axes (e.g., vertical and medial-lateral) (Cutti et al., 2010; Hafer et al., 2020; Lebleu et al., 2020; McGrath et al., 2018; Nazarahari and Rouhani, 2019). This procedure doesn’t give precise anatomical sagittal, frontal, or transverse axes. Walking-based functional orientation approaches can give accurate sagittal plane joint angles and eliminate the need for precise motions about each axis for orientation (Lebleu et al., 2020). Walking and standing are easy to capture in observed or real-world settings and thus lower extremity IMU data could be oriented to consistent vertical and medial-lateral axes whenever a participant walks, regardless of actual sensor location or orientation on a segment.

Previous studies have implemented functional sensor-to-segment orientation methods (Chardonnens et al., 2012; Fan et al., 2022; Favre et al., 2009; Nüesch et al., 2017; Öhberg et al., 2013; Pacher et al., 2022; Seel et al., 2014; Vargas-Valencia et al., 2016), generally with the assumption that these methods orient IMU data to a reliable reference frame. However, we do not know whether these sensor-to-segment orientation methods provide consistent segment kinematics across different sensor placements. Variations in sensor placement may exist between research groups but sensor placement may also vary slightly within research groups between participants or data collection sessions. In the most extreme cases, sensor placement would be expected to vary to some extent in studies where participants place sensors themselves and collect data in unobserved settings. Understanding the extent to which a functional sensor-to-segment orientation can correct for such variations will enable better evaluation of results between studies with different sensor placements and will allow for an understanding of the potential effects of inter-subject or inter-session variations in sensor placement on IMU-derived kinematics. Therefore, the purpose of this study was to determine to what extent a sensor-to-segment functional orientation method minimizes the effect of differences in IMU sensor placement on segment angular excursions during walking. Because body composition and gait characteristics change with age and might affect sensor motion characteristics, a secondary purpose was to determine whether age influences the effect of functional orientation.

## Methods

### Participants

Twenty healthy adults, 10 aged 21-35 years (5 male, 28.2±3.7 years) and 10 aged 55-70 years (5 male, 60.8±3.3years) participated in this study. All subjects had a BMI <30 kg/m^2^, were able to walk for 30 min without assistive devices, and had no history of major traumatic injury, surgery, or chronic pain in the back or lower extremities. All individuals completed IRB-approved informed consent before completing any study procedures.

### Data Collection

Three IMUs (Opal v2, APDM/Clario, Philadelphia, PA) were placed on the pelvis and right thigh, shank, and foot in <u>typical</u>, <u>shifted</u>, or <u>rotated</u> positions (Figure 1). Pelvis sensors were placed over the sacrum (<u>typical</u>), lateral to the sacrum (<u>shifted</u>), and medial to the right anterior superior iliac crest (<u>rotated</u>). Thigh and shank sensors were placed at the midpoint on the lateral aspect of the segment (<u>typical</u>), proximal to typical (<u>shifted</u>), and on the anterior aspect of the segment (<u>rotated</u>). Foot sensors were placed on participants’ shoes on the dorsum of the foot (<u>typical</u>), on the lateral instep (<u>shifted</u>), and on the heel (<u>rotated</u>). <u>Shifted</u> placements represent possible inter-subject or inter-session variability and <u>rotated</u> placements represent alternative sensor placements seen in previous studies (Anwary et al., 2018; Rampp et al., 2015; Rebula et al., 2013; Seel et al., 2014; Vargas-Valencia et al., 2016). IMU data (3-axis accelerometer and gyroscope signals) were collected continuously at 128 Hz via synchronized logging as participants completed functional tasks (walking and toe-touches) and walked overground at a self-selected preferred speed.

**Figure 1.**
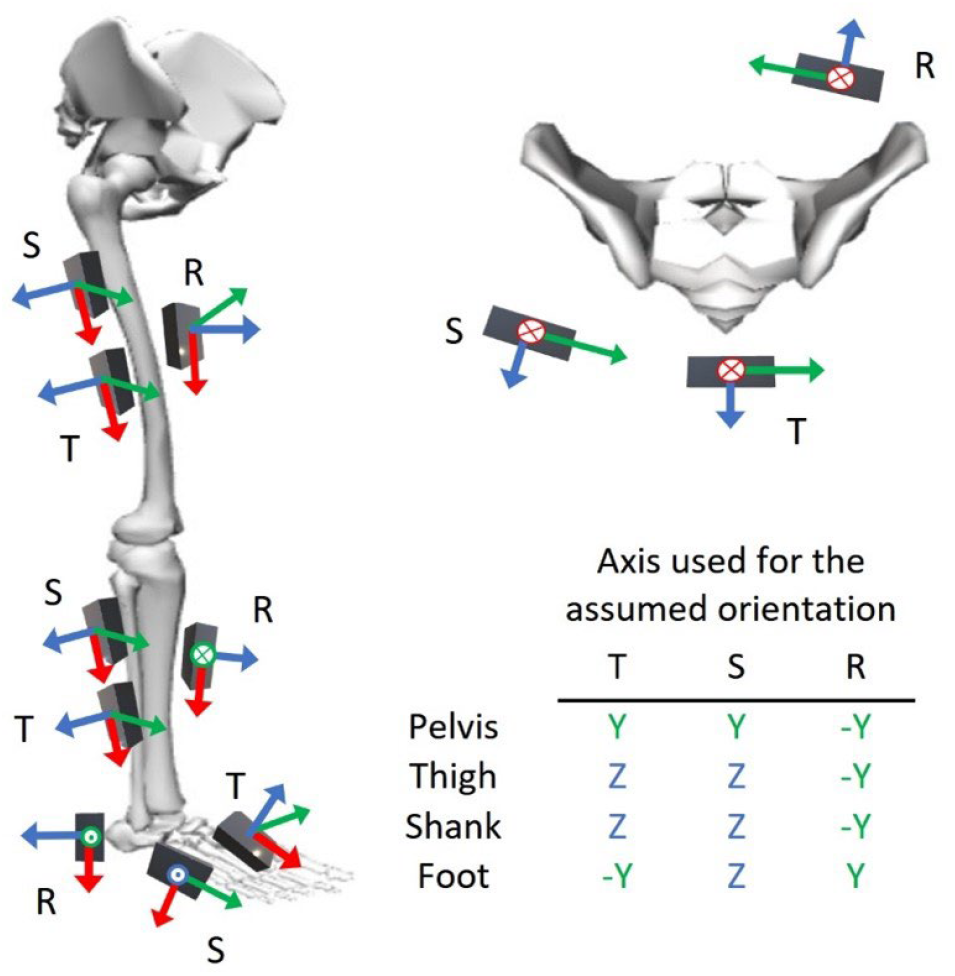
IMU placement with hardware sensor axes. Pelvis sensors were placed over the sacrum (typical, T), lateral to sacrum (shifted, S), and medial to the right anterior superior iliac crest (rotated, R). Thigh and shank sensors were placed at the midpoint on the lateral aspect of the segment (T), proximal to typical (S), and on the anterior aspect of the segment (R). Foot sensors were placed on the dorsum of the foot (T), on the lateral instep (S), and on the heel (R). X, Y, and Z-axes are red, green, and blue. Circles with an x indicate axis points directly into the page while dots point out of the page. Table presents the primary axis of rotation used for each sensor placement for the assumed orientation method.

### Data processing

#### Reference frame orientation

Data were processed via custom MATLAB scripts (R2019a, MathWorks, Natick, MA). Data processing began with identifying the data that included functional movements and postures: quiet standing, a short bout of straight-line walking, and toe touches (Figure 2). After functional data identification, we oriented IMU data for each sensor to a world reference frame, an assumed reference frame, and to two versions of a functional reference frame.

**Figure 2.**
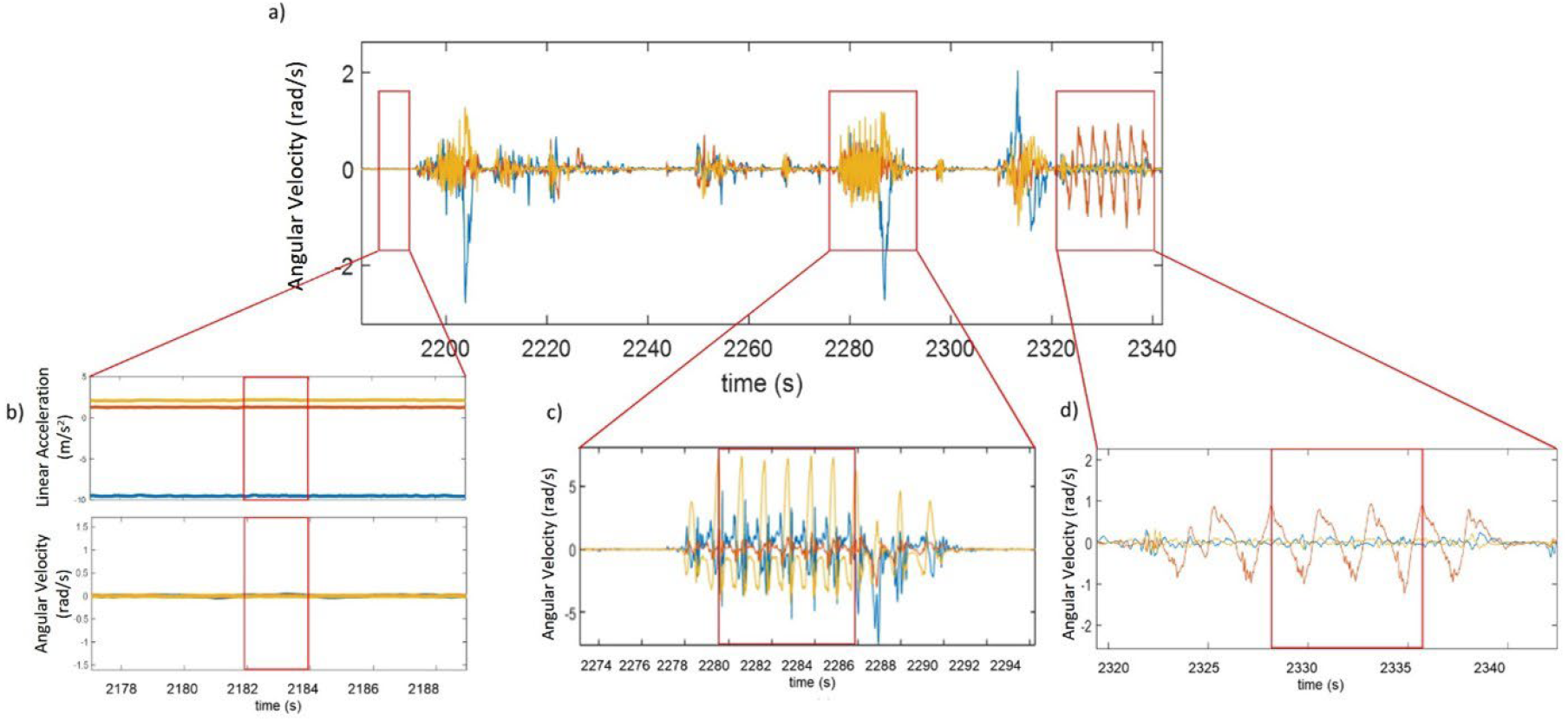
Calibration section of full data collection (a) including quiet static standing (b), walking (c), and toe touches (d). Functional tasks were identified using the typical pelvis and shank sensors’ linear acceleration and angular velocity data in the sensor hardware reference frames. a) Pelvis raw angular velocity data used to determine ranges of data for static standing, walking, and toe touches. b) A short range of pelvis raw linear acceleration (top) and angular velocity (bottom) data centered around zero (with gravity removed from acceleration signal) identified quiet standing. c) Several complete cycles of walking were selected based on peaks of shank raw angular velocity data and used to create the functional axes for the thigh, shank, and foot. d) Several complete cycles of toe touch cycles were selected based on peaks of pelvis raw angular velocity and used to create the functional axis for the pelvis.

##### World orientation

World orientation aligned the vertical (Z) axis with gravity and makes X and Y axes orthogonal in the horizontal plane (Figure 3). Orientation to the world reference frame was performed with a manufacturer-provided sensor fusion algorithm. World oriented data were only used for spatiotemporal variable calculations.

**Figure 3.**
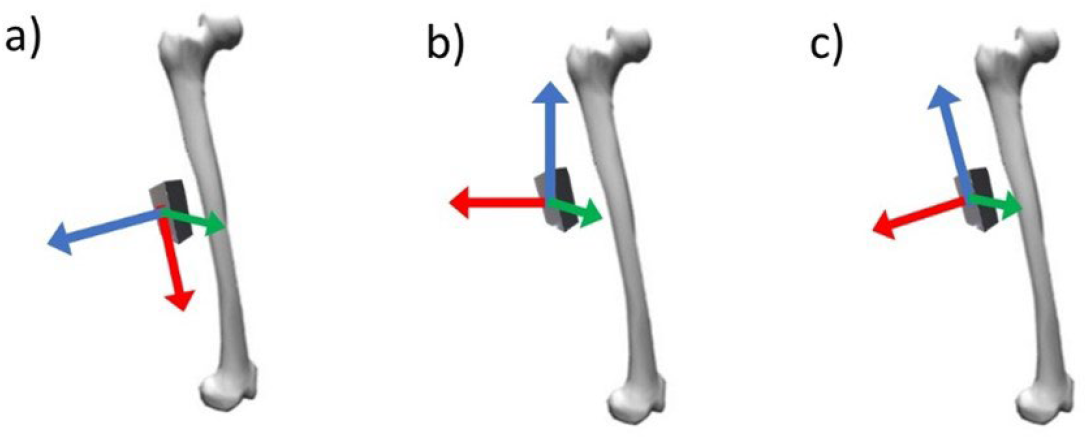
Example reference frame orientations. a) IMU hardware axes b) World oriented axes, Z aligned with gravity and X and Y orthogonal in the horizontal plane c) Functionally oriented axes, X and Z aligned by walking and quiet standing respectively.

##### Assumed sensor-to-segment orientation

Typical IMU placements represented where a researcher would place sensors such that specific sensor hardware axes are assumed to align with anatomy (e.g., for the thigh typical sensor placement, the +Z_sensor_ axis points laterally and so was assumed to align with the medial-lateral anatomical axis). For each sensor, the assumed axis was the hardware axis that aligned most closely with a segment’s medial-lateral anatomical axis (Figure 1).

##### Functional sensor-to-segment orientation

Initial functional reference frame axis orientations were defined by setting the longitudinal (Z_fun_) axis as parallel to gravity during static standing (i.e., longitudinal axis aligned with average linear acceleration vector). The medial-lateral (X_fun_) axis was the primary axis of rotation during straight-line walking (thigh, shank, foot) or toe-touches (pelvis). The primary axis of rotation was identified by passing each sensor’s raw angular velocity data from the functional movements through a principal component analysis (first principal component from pca function in MATLAB). An anterior-posterior (Y_fun_) axis was created as Z_fun_×X_fun_ (Figure 3c).

Because the initial longitudinal and medial-lateral axes are not necessarily perpendicular, one of these two axes must be re-oriented to ensure an orthogonal reference frame. Thus, there are two ways to define the functional reference frame. We created an <u>X-functional</u> reference frame holding X_fun_ constant while re-orienting the longitudinal axis as Z_x_fun_=X_fun_×Y_fun_. We also created a <u>Z-functional</u> reference frame holding Z_fun_ constant while re-orienting the medial-lateral axis as X_z_fun_=Y_fun_×Z_fun_.

### Walking identification

We extracted bouts of walking from continuous IMU data based on repeated oscillations of the shank sensor. Gait events were identified using a one-dimensional continuous wavelet transform procedure.

We used a ZUPT approach (Rebula et al., 2013) to calculate foot trajectories and stride lengths, velocities, and directions. See Supplement for more walking identification details.

This analysis only used straight-line steady-state strides. Straight-line steady-state was defined as strides with a consecutive inter-stride velocity difference of less than 0.1 m/s and angular deviation of less than 10°. Twenty-six strides were included per participant.

### Outcome variable calculation

Segment angular excursions about the medial-lateral axis were calculated for every stride for each sensor (typical, shifted, rotated) for assumed, X-functional, and Z-functional reference frames. Segment angular excursions were calculated by integrating the angular velocity between consecutive heel strikes. For the functional sensor-to-segment orientation methods, we integrated angular velocity about the medial-lateral (X) axis. For the assumed sensor-to-segment orientation method, we integrated angular velocity about the sensor axis that best aligned with the anatomical medial-lateral axis (Figure 1). The average segment angular excursion time series over 26 strides per orientation technique per IMU placement per subject were included in statistical comparisons.

### Statistics

For each sensor-to-segment orientation method (assumed, X-functional, Z-functional), segment excursion time series data were compared across each of the three IMU placements (typical, shifted, and rotated) and between groups (younger and older) using a continuous 2-way ANOVA via statistical parametric mapping (SPM; α=.05). Parametric and nonparametric implementations of the continuous 2-way ANOVA were run. We report parametric here as they agreed qualitatively with non-parametric results (non-parametric results reported in Supplementary files). Where significant interactions were found, independent t-tests via SPM were used to determine if excursions differed between placements within a group (Pataky, 2012). Where significant main effects of placement were found, we used t-tests via SPM to compare excursions between placements within an orientation method. RMSD was calculated between each of the three IMU placements (typical, shifted, and rotated) within each orientation method (assumed, X-functional, Z-functional) for all subjects to quantify the total difference in segment excursion time series between placements and sensor-to-segment orientation methods. We tested whether the mean and variance in RMSD between sensor placements differed using one-way ANOVAs and Bartlett’s tests, respectively.

## Results

There were no significant placement by group interactions for any segment. There was a significant main effect of sensor placement for the assumed sensor-to-segment orientation for all segment excursions except the foot (all p<0.001). There were also significant main effects of sensor placement for the X & Z functional orientations for the pelvis and thigh excursions (all p<0.001). All post-hoc findings reported in the following sections were significant at p<0.001 (Figure 4).

**Figure 4.**
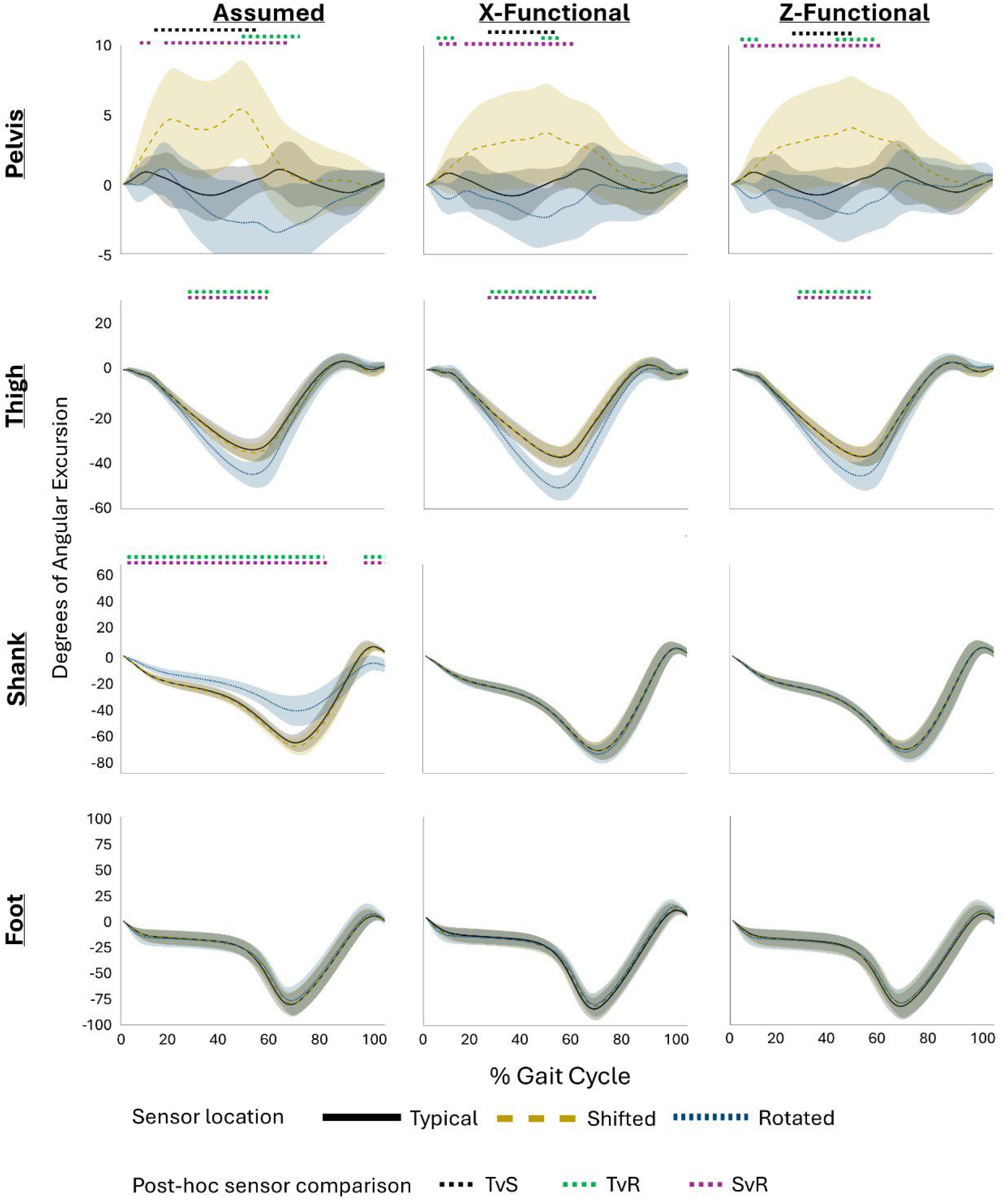
Segment excursions (mean and SD) for each sensor and orientation method. Solid black line = typical sensor location, yellow dashed line = shifted, and blue dotted line = rotated (see figure 1). Dashed lines above plots indicate significant post-hoc differences between sensor locations. Black = typical vs shifted, green = typical vs rotated, purple = shifted vs rotated.

### Assumed orientation

#### Main effects

For the assumed sensor-to-segment orientation, angular excursions differed between sensor placements at 8-68% of the gait cycle for the pelvis, at 26-57% of the gait cycle for the thigh, and at 2-78% and 92-100% of the gait cycle for the shank. Foot angular excursions did not differ between sensor placements.

#### Post-hoc comparisons

For the assumed orientation, pelvis angular excursions differed between typical and shifted (13-52% gait cycle), between typical and rotated (46-68% gait cycle), and between rotated and shifted placements (8-10% and 17-63% gait cycle). Thigh angular excursion differed between typical and rotated and between shifted and rotated placements (both 26-57% gait cycle). Shank angular excursion differed between typical and rotated (2-77% and 92-100% gait cycle) and between shifted and rotated placements (2-78% and 92-100% gait cycle).

### X-Functional orientation

#### Main effects

For the X-functional sensor-to-segment orientation, angular excursions differed between sensor placements at 5-12% and 17-61% of the gait cycle for the pelvis and at 28-69% of the gait cycle for the thigh. Shank and foot angular excursions did not differ between sensor placements.

#### Post-hoc comparisons

X-functionally oriented angular excursions for the pelvis differed between the typical and shifted (24-50% gait cycle), between typical and rotated (5-12% and 45-52% gait cycle), and between shifted and rotated placements (6-14% and 17-61% gait cycle). Thigh angular excursion differed between typical and rotated (29-66% gait cycle) and between shifted and rotated placements (28-69% gait cycle).

### Z-Functional orientation

#### Main effects

For the Z-functional sensor-to-segment orientation, angular excursions differed between sensor placements at 5-12% and 24-61% of the gait cycle for the pelvis and at 26-54% of the gait cycle for the thigh. Shank and foot angular excursions did not differ between sensor placements.

#### Post-hoc comparisons

Z-functionally oriented angular excursions for the pelvis differed between typical and shifted (24-52% gait cycle), between typical and rotated (5-12% and 45-57% gait cycle), and between shifted and rotated placements (6-61% gait cycle). Thigh angular excursion differed between typical and rotated (27-53% gait cycle) and between shifted and rotated placements (26-54% gait cycle).

##### RMSD

Mean subject RMSD between pelvis sensor placements ranged from 2.6-4.9° for assumed, 2.1-3.8° for X-functional, and 1.9-3.8° for Z-functional orientations. Mean subject RMSD between thigh sensor placements ranged from 1.4-5.7° for assumed, 1.5-7.7° for the X-functional, and 0.9-4.8° for Z-functional orientations. Mean subject RMSD between shank sensors ranged from 1.9-15.1° for assumed, 0.6-1.5° for X-functional, and 0.6-1.5° for Z-functional orientations. Mean subject RMSD between foot sensors ranged from 2.7-4.6° for assumed, 1.9-3.7° for X-functional, and 1.9-3.7° for Z-functional orientations. RMSD between sensor placements for each participant are shown in Figure 5.

**Figure 5.**
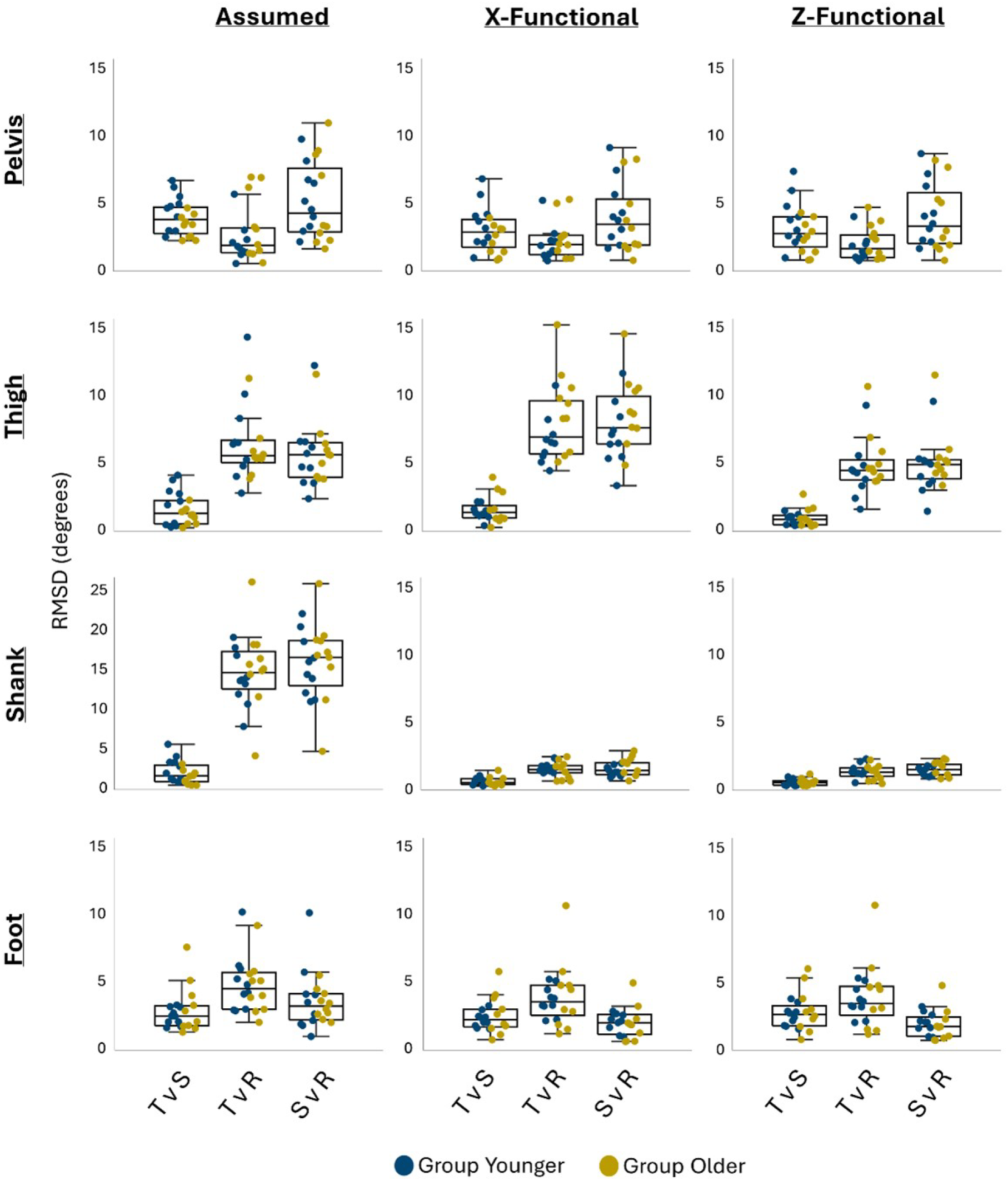
Root mean squared differences between sensor locations (T = typical, S = shifted, R = rotated) within orientation methods. Dots indicate individual subjects in the younger (blue) & older (yellow) groups. See figure 1 for illustration of sensor locations.

The RMSD for the pelvis excursions were not significantly different between sensor placements or orientation methods (all p>0.05). The RMSDs between typical and rotated thigh sensor placements and shifted and rotated thigh sensor placements were greater for the X-functional compared to assumed and Z-functional orientations (p<0.001). The RMSDs between all shank sensor placements and between typical and rotated foot sensor placements were greater for the assumed compared to the X and Z-functional orientations (p<0.001).

The variances of RMSD between typical and rotated pelvis sensor placements (p=0.04), between typical and shifted thigh sensor placements (p=0.04), between all shank sensor placements (p<0.001), and between shifted and rotated foot sensor placements (p=0.003) were greater for the assumed compared to the X and Z-functional orientations.

## Discussion

The primary aim of this project was to determine the extent to which a walking- and toe-touch-based sensor-to-segment functional orientation method minimizes the effect of differences in IMU sensor placement on segment angular excursions during walking. The functional sensor-to-segment orientation method reduced differences in angular excursion between IMU placements, particularly for the pelvis and shank. While differences in angular excursion for the assumed sensor-to-segment orientation method encompassed up to 60% of the gait cycle for the pelvis, 31% for the thigh, and 84% for the shank, functional orientation reduced these ranges to 44% of the gait cycle for the pelvis, 28% for the thigh, and 0% for the shank. These trends did not differ with age. These findings suggest that this functional sensor-to-segment orientation method can minimize differences in segment excursion caused by variations in sensor placement, unless sensors are placed on areas with substantial soft tissue artifact.

X-& Z-functional sensor-to-segment orientation methods similarly minimized inter-placement differences in segment excursion compared to the assumed sensor-to-segment orientation, with Z-functional performing slightly better. The effect of the functional orientation method is best demonstrated by the similarity of the data between all sensor placements for the shank (Figures 4 & 5). The shank had the largest range of significantly different assumed orientation angular excursions across sensor placements but had no differences in angular excursion between sensors after functional orientation. This may be because the shank resembles a rigid body, making the true angular velocity magnitude similar regardless of sensor placement on the segment (Scalera et al., 2021).

The X-& Z-functional orientation methods performed similarly for the thigh and pelvis, but with larger ranges of differences between sensor placements compared to the shank and foot. These differences may be due to the effect of soft tissue artifact on the thigh and pelvis being larger than that at the shank or foot. For the rotated thigh placement, soft tissue artifact is likely adding motion about the medial-lateral axis, thus increasing the segment excursion about the primary axis of rotation (Stagni et al., 2005). This can be seen where the greatest difference between sensor placements occurred during mid to late stance (Figure 4) when the quadriceps contract to produce knee extension. The shifted pelvis sensor’s motion could have been influenced by deformation of the gluteal muscles and overlying soft tissue during walking and soft tissue artifact during toe-touches may have affected the orientation of the functional axis for the rotated pelvis sensor. This hypothesis is supported by previous work that found that hip angles were most sensitive to sensor placement during activities with increased flexion angles (Horenstein et al., 2019). These thigh and pelvis findings support a recommendation that sensors should be placed away from areas that are subject to deformation from muscle or soft tissue.

The RMSD results allow us to evaluate the overall difference in segment excursions between two sensor placements within an orientation method. Smaller compared to larger RMSDs indicate that excursions from two placements are more similar. When compared across orientation methods, the method with the smaller RMSD produces more similar segment excursion estimates regardless of sensor placement. In this study, RMSD between sensor placements was smaller for both functional sensor-to-segment orientation methods compared to assumed sensor-to-segment orientation for all comparisons except the pelvis X- and Z-functional orientations and the thigh X-functional orientation. Additionally, the mean RMSDs for all but 2 comparisons were smaller than 5°, which is generally considered the threshold for a clinically meaningful difference in joint angles (McGinley et al., 2009). Variance in RMSD between sensor placements was also smaller for both X and Z-functional orientations compared to the assumed orientation for the pelvis typical vs rotated placements, thigh typical vs shifted placements, for all shank placements, and for the foot shifted vs rotated placements. Smaller RMSD variance supports the robustness of the orientation method to minimize the effect of placement across all participants.

Although the functional sensor-to-segment orientation methods produced lower RMSDs compared to the assumed sensor-to-segment orientation method overall, there were a few outliers. The largest RMSD for both thigh functional orientation segment excursions in the older group was a female with the highest body mass. This finding aligns with our hypothesis that soft tissue artifact may add angular velocity signal about the medial-lateral axis during gait. The highest RMSD for the thigh Z-functional orientation in the younger group was the tallest male with the lowest BMI. This participant was an avid runner. His low BMI and more developed quadriceps may have led to increased sensor movement about the medial-lateral axis during gait due to muscle deformation. Finally, the highest RMSD for both foot functional orientation segment excursions was the shortest older female. We hypothesize that the smaller surface area for sensor placement on her feet may have led to an increased effect of the metatarsophalangeal joint on sensor movement about the medial-lateral axis. These effects of anthropometrics were only seen when comparing typical and shifted to rotated sensor placements, further supporting the recommendation of avoiding sensor placement on areas that experience non rigid-body motion.

In this study, IMU placements represented possible inter-subject, inter-session, or inter-research group differences in sensor placements, as well as possible differences in sensor placement in unobserved, real-world data collection. In practice, small differences in sensor placement can be hard to identify when processing data or comparing across studies. The rotated position is an extreme example, but our findings that the functional sensor-to-segment orientation method was still able to minimize many of the effects of this placement implies that the effect of smaller differences in sensor placement would be effectively minimized by functional sensor-to-segment orientation, if large areas of soft tissue artifact are avoided.

Demonstrating the extent to which a functional sensor-to-segment orientation method minimizes the effect of varying sensor placement is an important step towards comparing across studies with similar data orientation methods and collecting reliable inter-subject, inter-session, and real-world data. The demonstrated walking- and toe-touch-based functional orientation method can correct for shifts in sensor placement, if sensors are not placed on areas of large soft tissue deformation. This relatively easy-to-implement sensor-to-segment orientation method is a promising approach to improve between-study comparability of IMU-derived gait mechanics as well as decrease inter-subject and inter-session variability within studies. Additionally, this sensor-to-segment orientation method may be an effective means of ensuring reliable estimation of segment kinematics in unobserved, free-living data collections where sensor placement may vary more than in controlled lab settings both within and between data collection sessions.

## Data Availability

All data produced in the present study are available upon reasonable request to the authors

## Acknowledgements

This research was supported by the National Institutes of Health’s National Institute on Aging, grant R21AG076989

## Supplement: Walking identification, gait event identification, and spatiotemporal calculation details; Non-parametric statistical results

### Walking identification

We extracted bouts of walking from continuous IMU data based on repeated oscillations of the shank sensor (Figure 1S). First, for consecutive five-second windows of data, data from each axis of the shank sensor raw angular velocity signal (Figure 1S a.) was passed through a fast Fourier transform (Figure 1S b.). Then, within each window, the frequency power from all three axes were summed to determine the total power density. Windows with high-power density (power greater than 1/3 the maximum power) within frequencies representative of shank oscillations during walking (0.5-2.2 Hz) were considered to contain walking data. Consecutive identified sections of walking combined (Figure 1S c.).

### Gait event identification

Individual strides were identified using data from the foot sensor. Foot world-oriented vertical acceleration data were passed through a one-dimensional continuous wavelet transform (Figure 1 Sd.). The absolute value of the first (i.e., highest-frequency) wavelet was low-pass filtered with a second order Butterworth filter with a cut-off frequency of 4Hz. Signal peaks above a threshold (half the median magnitude of the peaks of the filtered signal) were defined as gait events. To determine heel strike and toe off we first found periods of foot flat using a zero-velocity update algorithm on the foot world frame data using acceleration and velocity magnitude thresholds of 1 m/s^2^ and 1 rad/s, respectively. We then integrated the foot world frame acceleration signal between periods of foot flat to find foot velocity and displacement. Heel strike and toe off were defined using the slope of the vertical foot sensor displacement at the time of each identified gait event. A negative slope indicated heel strike and positive slope indicated toe off. A stride was created if gait events included a consecutive heel strike - toe off - heel strike pattern.

### Spatiotemporal calculations

Stride displacement was calculated as the double integral of the typical foot sensor linear acceleration in the world reference frame. Stride length was the net displacement in the horizontal direction between heel strikes. Stride velocity was calculated by dividing stride length by the time between heel strikes. Stride angular deviation was calculated by taking the inverse tangent of the displacement in the two horizontal world reference axis directions.

**Figure 1S.**
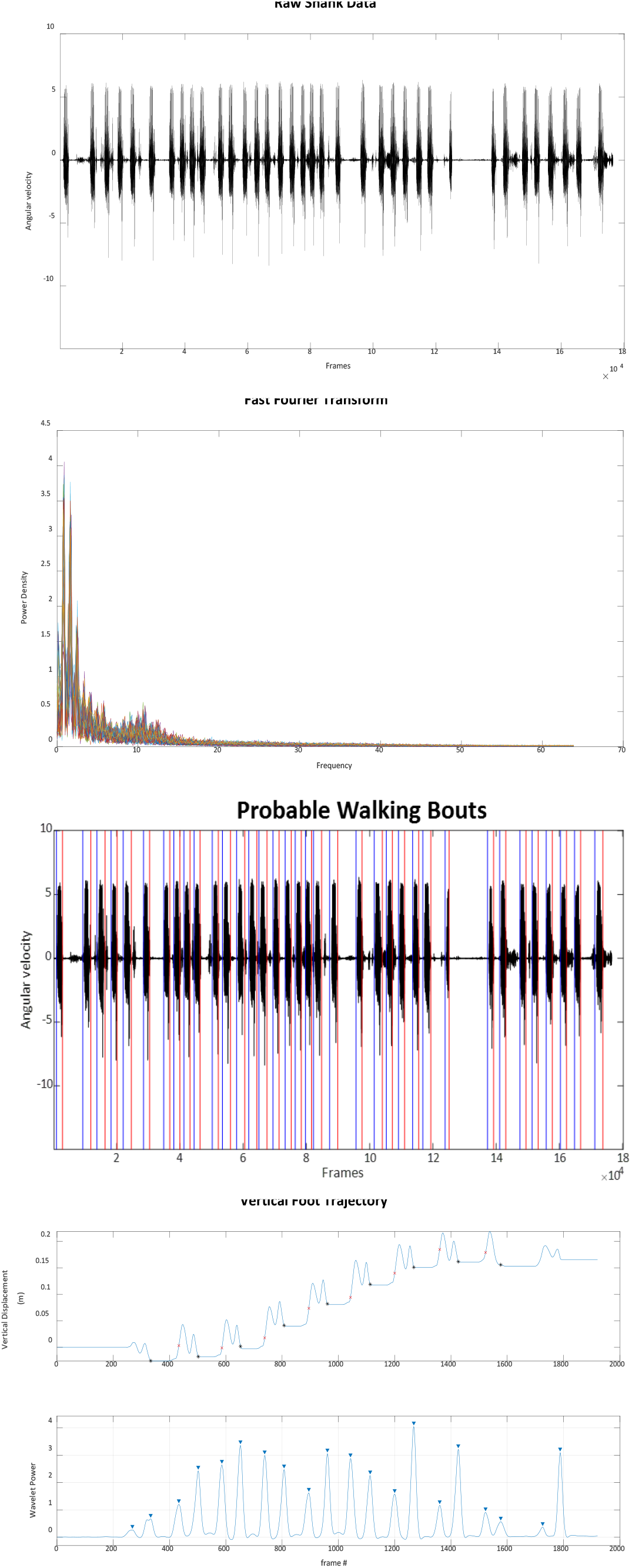
Walking and gait event identification procedure. a) Range of raw shank angular velocity data that may contain walking. b) Each line represents one Fast Fourier transform of one 5 second window. c) Identified ranges of walking based on windows with a high-power density and of a frequency between 0.5 and 2.2 Hz. d) Top plot visualizes the foot vertical displacement. Black * represents heel strikes and red x represents toe offs.

### Nonparametric SPM angular excursion time series results

#### Assumed orientation

##### Main effects

For the assumed sensor-to-segment orientation, pelvis angular excursions differed between sensor placements at 5-75% of the gait cycle. Thigh angular excursions differed between sensor placements at 24-58% of the gait cycle. Shank angular excursions differed between sensor placements at 2-80% and 92-100% of the gait cycle. Foot angular excursions did not differ between sensor placements.

#### X-Functional orientation

##### Main effects

For the X-functional sensor-to-segment orientation, pelvis angular excursions differed between sensor placements at 2-61% of the gait cycle. Thigh angular excursions differed between sensor placements at 28-74% of the gait cycle. Shank and foot angular excursions did not differ between sensor placements.

#### Z-Functional orientation

##### Main effects

For the Z-functional sensor-to-segment orientation, pelvis angular excursions differed between sensor placements at 5-61% of the gait cycle. Thigh angular excursions differed between sensor placements at 24-54% of the gait cycle. Shank and foot angular excursions did not differ between sensor placements.

